# International healthcare experts’ consensus on the key requirements of a potential international patient safety learning system: a modified online Delphi study

**DOI:** 10.64898/2026.02.25.26347126

**Authors:** Jaafer Qasem, Adrian Edwards, Fiona Wood, Andrew Carson-Stevens

## Abstract

**Background:** Despite widespread recognition that patient safety learning can transcend national boundaries, no international patient safety learning system (PSLS) currently exists. There is no expert consensus on the purpose, key requirements, or feasibility of such a system.

**Objective:** To gain consensus from an international panel of healthcare experts regarding the key requirements and feasibility of a potential international PSLS, with or without an incident reporting function.

**Methods:** A two-round modified online Delphi study was conducted with 21 international healthcare experts in patient safety and learning systems, representing all six continents. The study was informed by a prior systematic literature review and semi-structured key-informant interviews with safety-critical industry experts. Panellists rated statements on a 9-point Likert-like scale. Consensus was defined a priori as ≥70% agreement (ratings 7–9) with an interquartile range (IQR) ≤2.00. A post-hoc threshold of ≥80% was applied to identify the strongest areas of consensus.

**Results:** Of 73 experts invited, 21 completed round one (29% response rate) and 15 completed round two (71% retention). Across two rounds, 103 statements were evaluated; consensus was achieved on 85 (83%) at the ≥80% threshold across all four domains: purposes (15/19 statements); key functions and features (17/22); patient safety incidents and criteria for international concern (19/24 combined); and enablers and challenges (34/38). The panel endorsed a proposed framework for an international PSLS and generated novel consensus-based lists of patient safety incidents and criteria for determining what should be shared internationally.

**Conclusions:** International healthcare experts broadly agree on the purposes, key functions, and feasibility of an international PSLS. The consensus-derived framework provides a foundation for future feasibility research and potential pilot implementation. Validation with frontline end-users is an essential next step.

**KEY MESSAGES:** *What is already known on this topic:* National patient safety learning systems vary considerably in design, governance, and the degree to which they generate actionable learning, and no international system currently exists to enable systematic cross-border sharing and learning from patient safety data.

*What this study adds:* This is the first Delphi study to establish international expert consensus on the purposes, key functions and features, and feasibility of an international Patient Safety Learning System (PSLS), producing novel consensus-based lists of patient safety incidents relevant to international sharing and criteria for determining what constitutes a risk of international concern.

*How this study might affect research, practice or policy:* The proposed framework and consensus-derived criteria provide a starting point for feasibility research and potential pilot implementation by organisations such as the World Health Organization (WHO), in alignment with the Global Patient Safety Action Plan 2021–2030. The findings also highlight the structural prerequisites — including a standardised international taxonomy, governance frameworks, and equitable participation mechanisms — that must be addressed before implementation can proceed.

## INTRODUCTION

Patient safety incidents remain a significant source of morbidity and mortality across healthcare systems worldwide.^[1–4]^ Since the publication of seminal reports on the inadvertent effects of healthcare provision, there has been growing recognition of the need for systematic strategies to monitor adverse events and learn from safety data.^[1,2]^ While national and local patient safety learning systems (PSLSs) have been established in many countries to facilitate the reporting, analysis, and learning from such incidents,^[6]^ these systems vary considerably in design, governance, and the degree to which they generate actionable learning.^[7,8]^ No international PSLS currently exists to enable the systematic sharing and learning from patient safety data across national boundaries.

The potential value of international learning in patient safety has been recognised by the WHO, which has identified the need for collaborative strategies to promote the dissemination of learning across countries.^[3–5]^ The WHO Global Patient Safety Action Plan 2021–2030 explicitly calls for mechanisms to facilitate international sharing and learning from patient safety events.^[5]^ Despite this recognition, existing international initiatives — such as pharmacovigilance networks and WHO Collaborating Centres — address only specific aspects of patient safety and often lack the capacity to support broader, routine cross-national learning.^[9–11]^ The heterogeneity in how national systems collect, classify, and analyse patient safety data presents a further structural challenge.^[7,10]^

A prior systematic literature review conducted by the authors identified the key features and functions that might be required for an international PSLS, drawing on evidence from both healthcare and other safety-critical industries such as aviation and nuclear power.^[14]^ This was followed by semi-structured key-informant interviews with eleven experts from safety-critical industries, which further explored the requirements, enablers, and challenges of such a system.^[15]^ Together, these studies identified four key themes requiring further exploration: (1) the purpose of an international PSLS; (2) the operationalisation of that purpose through functions and features; (3) the identification and sharing of internationally relevant patient safety incidents; and (4) enablers and challenges to establishing such a system. The synthesis generated a preliminary framework for an international PSLS, along with unanswered questions requiring expert consensus. As the evidence base was largely derived from safety-critical industries outside healthcare, validation by healthcare-specific experts was essential to determine the transferability of these findings.

This study aimed to gain consensus from a broad panel of international healthcare experts regarding the key elements required for a potential international PSLS, with or without an incident reporting function. Specific objectives included gaining consensus on: the purposes and key requirements of an international PSLS; its feasibility in terms of enablers and challenges; a list of patient safety incidents deemed relevant for international sharing and learning; and a set of criteria for determining whether a patient safety risk is of international concern.

## METHODS

### Study design

A two-round modified online Delphi study was conducted between June and November 2020. The study was informed by a multimethod, multiphase approach in which a systematic literature review^[14]^ and semi-structured key informant interviews^[15]^ replaced the traditional first qualitative round of the Delphi process.^[16–18]^ The Delphi technique was selected for its capacity to facilitate anonymous, structured consensus-building among geographically dispersed experts.^[18–21]^ Ethical approval was obtained from Cardiff University’s School of Medicine Research Ethics Committee (SMREC Reference Number: 18/70).

### Expert selection and recruitment

Experts were defined as individuals with knowledge and experience in patient safety, incident reporting or learning systems, organisational learning, or healthcare services, consistent with established definitions.^[24,30]^ Experts were identified through: authorship of studies included in the systematic review (n=18); prior inclusion in the interview study (n=9); participation in WHO consultative meetings on patient safety (n=44); patient group representation (n=2); and frontline healthcare staff with relevant expertise (n=6). Snowball sampling yielded an additional three participants.^[33]^ All experts met predefined inclusion criteria (online supplementary table S1). A minimum panel size of 20 was targeted, consistent with published Delphi studies of similar scope.^[20,34,35]^ The expert identification and recruitment process is illustrated in figure 1.

**Figure 1.**
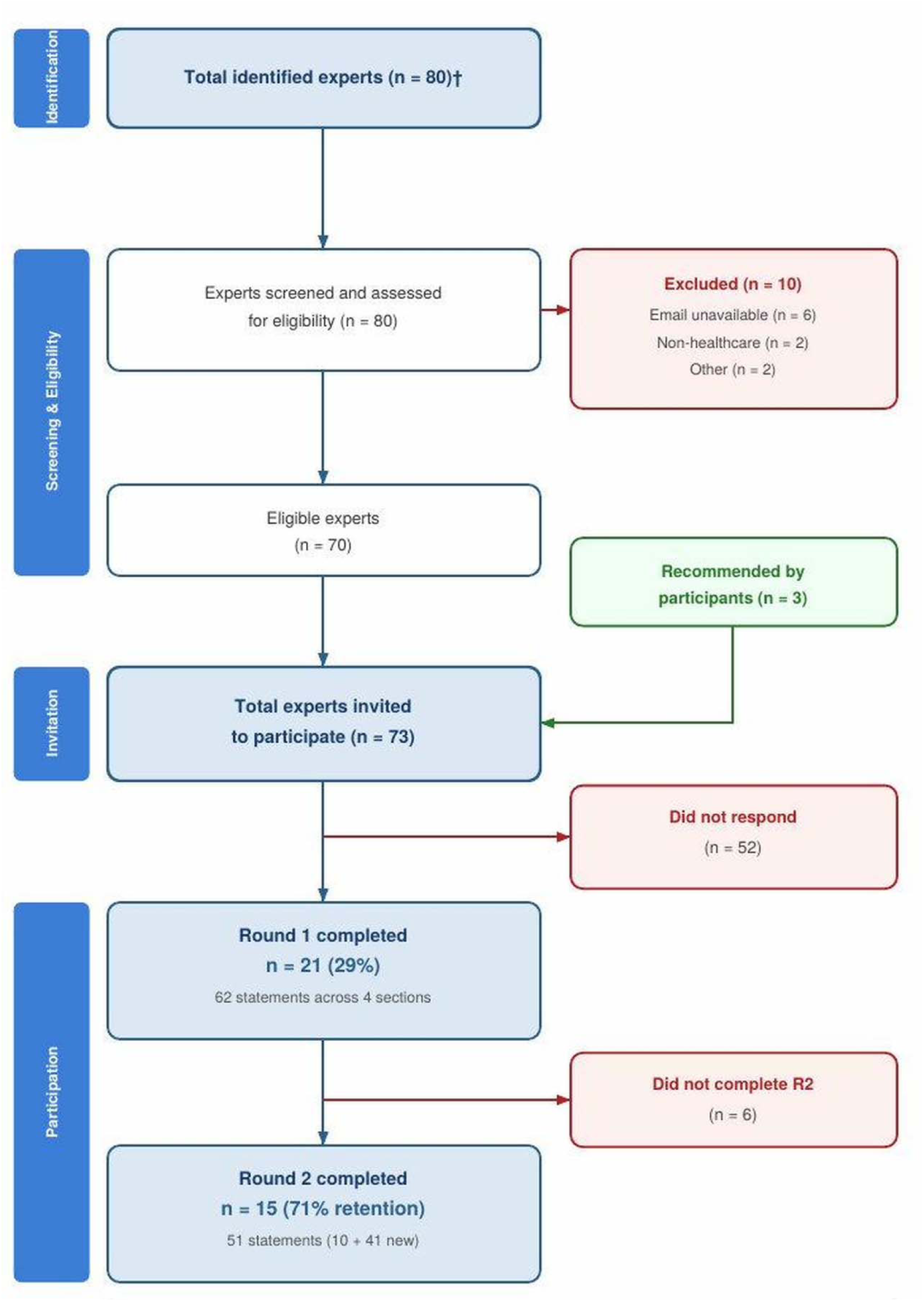
The process of identification and recruitment of experts for the Delphi panel.

### Survey development

Delphi statements were developed through triangulation of the systematic review and interview findings, producing an integrated set across four thematic sections: (1) purposes of an international PSLS; (2) key functions and features; (3) patient safety incidents relevant to international sharing and learning; and (4) enablers and challenges. Statements were refined iteratively within the research team and pilot tested with two subject matter experts external to the panel. Pilot testing led to modifications including an electronic consent form, reduction in estimated completion time from 40 to 30 minutes, and refinement of statement wording for clarity. The survey instrument was further validated by experts at a WHO Collaborating Centre in Human Factors and Communication for the Delivery of Safe and Quality Care in Florence, Italy. The survey was administered electronically via the Qualtrics platform (Qualtrics, Provo, UT, USA).

### Delphi procedure

In round one, panellists rated their agreement with 62 statements on a 9-point Likert-like scale (1=strongly disagree; 9=strongly agree).^[18,21,22]^ Justifications were requested for ratings of 6 or below, and optional free-text feedback was invited after each section.^[17,40]^ In round two, panellists who completed round one received de-identified group results (median, percentage agreement, interquartile range [IQR]) for each statement, and were asked to re-rate statements that had not reached consensus and to rate newly generated statements derived from qualitative feedback of round one. New statements underwent the same iterative refinement process within the research team before inclusion.

### Consensus definition and termination criteria

Consensus was defined a priori as ≥70% agreement (ratings 7–9) with an IQR ≤2.00, in line with recommended quality indicators for Delphi studies.^[16,25,39]^ The Delphi was to be terminated if the panel reached consensus on ≥80% of all statements across both rounds, or if the response rate fell below 70% in the second round.^[16,18]^ Given that a large proportion of statements reached consensus, the threshold was raised post-hoc from 70% to 80% agreement to identify the strongest areas of consensus and to produce a more focused set of recommendations. This adjustment is acknowledged as a study limitation.

### Data analysis

Percentage agreement, median, and IQR were calculated for each statement.^[25,26]^ Qualitative feedback was reviewed thematically to generate new statements for round two. Statements reaching consensus were ranked within each section by percentage agreement, IQR, and median.

## RESULTS

### Panel characteristics

Of 73 experts invited, 21 completed round one (29% response rate; figure 1). The panel comprised medical doctors (n=8), researchers (n=5), academics (n=3), healthcare leaders/managers (n=3), a patient representative (n=1), and a medical coder (n=1), with a combined total of 314 years of experience in patient safety (mean 14 years; range 3–32 years). Panellists represented all six continents (figure 2); full demographics are presented in table 2. In round two, 15 of 21 panellists responded (71% retention).

**Figure 2.**
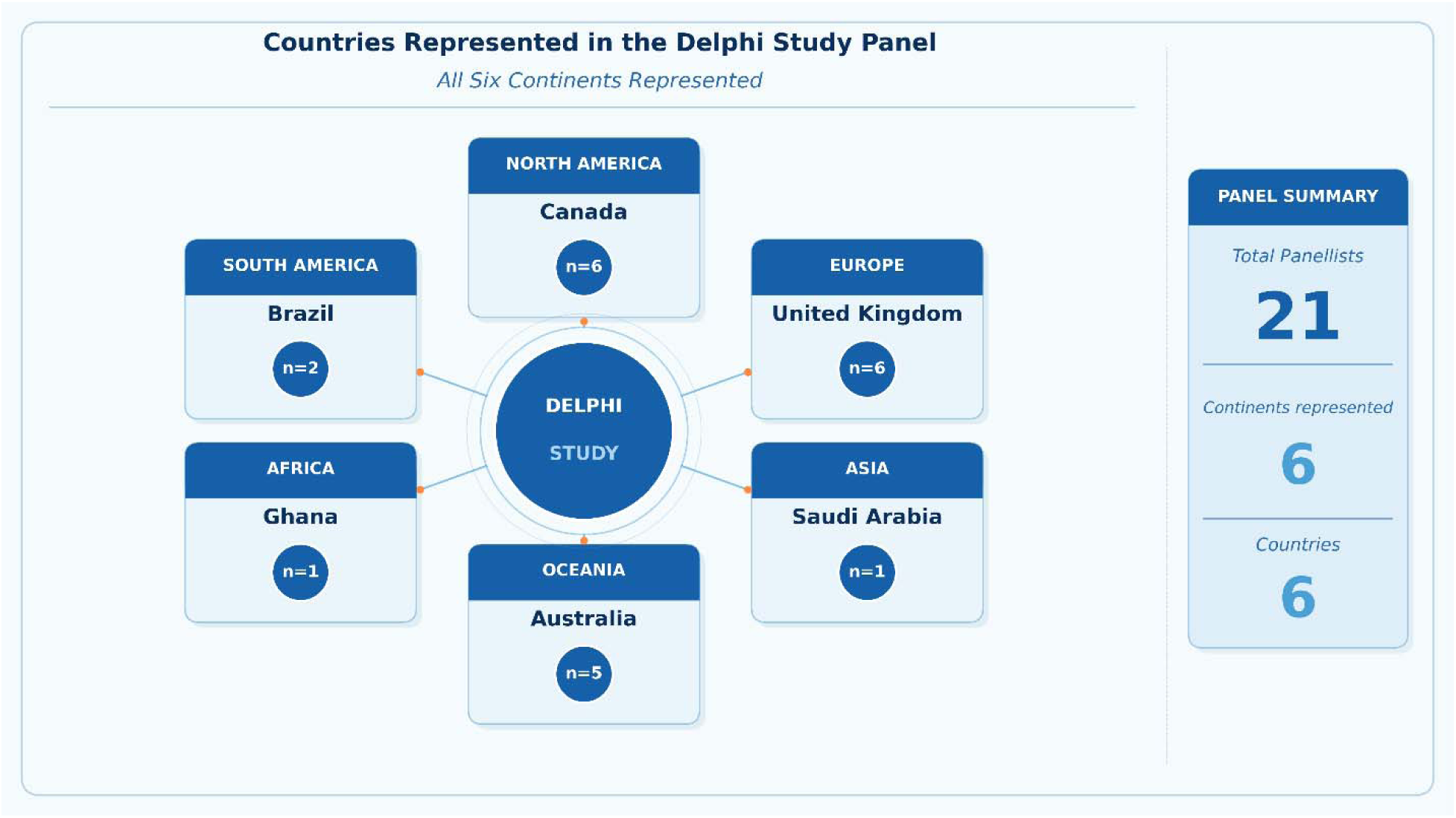
Countries represented in the Delphi panel and the number of panellists per country.

### Overview of consensus

Round one presented 62 statements across four sections; consensus was achieved for 52 (84%). Qualitative feedback generated 41 new statements for round two, including a novel subsection on criteria for determining what constitutes an internationally relevant patient safety risk. Round two presented 51 statements (10 carried forward; 41 new); consensus was achieved for 49 (96%). Across both rounds, the panel reached consensus on 101 of 103 statements (98%) at the ≥70% threshold and on 85 of 103 (83%) at the post-hoc ≥80% threshold. Table 1 summarises consensus rates by section. Full results for all 103 statements are provided in online supplementary table S2.

**Table 1.** Summary of consensus results by section (≥80% agreement threshold).

| Section | Statements (n) | Consensus $\geq 80\%$ (n) | Consensus rate | Non-consensus (n) |
| --- | --- | --- | --- | --- |
| 1. Purposes of an international PSLS | 19 | 15 | 79% | 4 |
| 2. Key functions and features | 22 | 17 | 77% | 5 |
| 3A. Incidents for international sharing | 16 | 11 | 69% | 5 |
| 3B. Criteria for international concern | 8 | 8 | 100% | 0 |
| 4A. Enablers | 23 | 19 | 83% | 4 |
| 4B. Challenges/barriers | 15 | 15 | 100% | 0 |
| **Total** | **103** | **85** | **83%** | **18** |

**Table 2.** Characteristics of the international Delphi expert panel.

| Profession | n | Mean years' experience (range) | Example of experience at national/international level | Example of leadership roles |
| --- | --- | --- | --- | --- |
| Medical doctor* | 8 | 12 (5–32) | Contributor to WHO medication safety initiatives | Chief quality and safety officer at university hospital |
| Researcher | 5 | 15 (8–28) | Consulted to the WHO | Senior executive in a national patient safety agency |
| Academic | 3 | 28 (20–32) | Government committees, international advisory work | Former Chief Medical Adviser for a Health Agency |
| Leadership/Management | 3 | 15 (10–20) | Director of a national patient safety centre | Executive leader of national agency |
| Patient representative | 1 | 12 | Advisor to national and international patient safety research initiatives | Patient safety teaching in medical and nursing programmes |
| Medical coder | 1 | 3 | Works for a national patient safety centre | N/A |

*\*Including physician and medical specialist (e.g., cardiologist).*

### Purposes of an international PSLS

The panel endorsed a broad set of purposes for an international PSLS. The strongest consensus was on the identification of research and development priorities to focus international efforts (100% agreement, median 8), the identification of evidence–practice gaps occurring in similar contexts across countries (100%), and the sharing of affordable design-based interventions to reduce patient safety risks (100%). The panel also strongly agreed that the system should facilitate learning with and from countries about what works and how (93%) and process learning from investigations to identify transferable lessons (90%). Notably, the panel endorsed both reactive functions (learning from incidents that have occurred) and proactive functions (surveillance to detect emerging risks), reflecting the conceptual distinction between a learning system and a purely incident reporting system. Four statements did not reach the ≥80% threshold: surveillance of patient safety incidents to detect potential risks internationally (76%), learning from patients and families about risk, harm, response, and remediation (73%), coordinating the design of initiatives/interventions to mitigate commonly identified patient safety risk (73%), and standardising the way learning from investigating patient safety incidents is reported (67%).

### Key functions and features

The highest-ranked statement was the development of a structured process for investigating patient safety risks identified by the international system (100% agreement). Experts strongly agreed that in cases of equipment- or medication-related incidents, manufacturers should be informed to take action (95%), and that safety recommendations with transferable solutions should be generated and shared (95%). Development of methodological approaches for assimilating descriptive patient safety data across systems (93%) and supporting countries’ progression towards proactive clinical risk management (93%) also received strong consensus. Five statements did not reach the ≥80% threshold, all at 76% agreement: supporting improvement in incident reporting and learning through sharing exemplars, the ability to share reported patient safety incidents/risks relevant to international learning, the ability to analyse and make recommendations based on shared patient safety data to identify transferable learning, the ability to trigger an alarm deploying notifications to countries about serious identified patient safety risks, and supporting improvements in the collection of patient safety data through sharing exemplars. The clustering of these five statements at the same agreement level suggests a degree of panel uncertainty around the more operational and data-sharing functions of the system, in contrast to the stronger consensus on its governance and learning-oriented functions.

### Patient safety incidents relevant to international sharing

Incidents involving products or devices as major contributing factors (100%), incidents where risk of severe harm or death is likely upon recurrence (100%), and drug and equipment safety incidents (100%) were rated as most relevant for international sharing. Patient safety risks new to a given country received strong consensus (93%), as did diagnostic error-related incidents (93%). Incidents related to manufacturing and supply chain issues (95%), such as contaminated vaccines or intravenous fluids, and incidents involving faulty medical devices (95%) were also strongly endorsed. Five statements did not reach the ≥80% threshold: ten-times medication errors (76%), tubing misconnection errors (76%), nasogastric tube positioning errors (76%), incidents resulting from relatively novel contributory factors such as pandemics, civil unrest, or natural disasters (73%), and any incident type impacting paediatric patients (73%).

### Criteria for international concern

The panel achieved 100% consensus on all eight statements in this section. The strongest endorsement was for the risk of harming large numbers of individuals across multiple countries if no intervention is taken (100%, median 9), followed by the morbidity and mortality impact of patient safety events (100%) and the need for international action from major stakeholders such as pharmaceutical companies (100%). The relevance of an identified risk to more than one country facing similar clinical challenges - for example, prevention and control of infectious disease - received 93% agreement.

### Enablers and challenges

Having a standardised international patient safety taxonomy received the highest consensus among enablers (100%, median 9), followed by the patient experience as a central facilitator for defining harm and setting mitigation priorities (100%). Easy and secure access for users and contributors (95%), buy-in from healthcare systems within individual countries (95%), representation from submitting countries (95%), and open accessibility of system outputs for learning (95%) were also strongly endorsed. Political will to generate and coordinate patient safety interventions of international concern received 93% agreement. Four statements did not reach the ≥80% threshold: availability of multiple input methods such as online, phone, and email (76%), password-controlled access to an online platform (73%), funding by partner countries (71%), and limiting access for input to one named responsible organisation per country (53%). The notably lower agreement on the last statement suggests the panel was divided on whether a centralised national gatekeeping model is desirable.

All 15 challenge/barrier statements reached consensus at the ≥80% threshold. The most strongly endorsed barriers were difficulty with funding (100%), potential establishment and maintenance costs (100%), lack of a data governance strategy (100%), and the value-proposition problem across varied jurisdictions (100%). The challenge posed by countries being at very different maturity levels with respect to safety culture received 87% agreement, and concerns about national reputation as a barrier to open data sharing reached 90% agreement.

### Proposed framework

Based on the consensus findings, combined with the prior systematic review and interview data, a framework for the design and operationalisation of an international PSLS was developed (figure 3). The framework conceptualises how national PSLSs would contribute patient safety data to a central international system, which would aggregate information, generate safety recommendations and alerts, and provide structured feedback to national systems and relevant stakeholders. For instance, identification of a contamination event in a batch of medication within any national system would feed into the international database, triggering analysis and generation of safety alerts issued to participating countries and feedback provided to manufacturers — such as the European Medicines Agency or national regulatory equivalents — to enact safety changes, while also enabling countries to learn from the experiences and interventions of others facing similar challenges.

**Figure 3.**
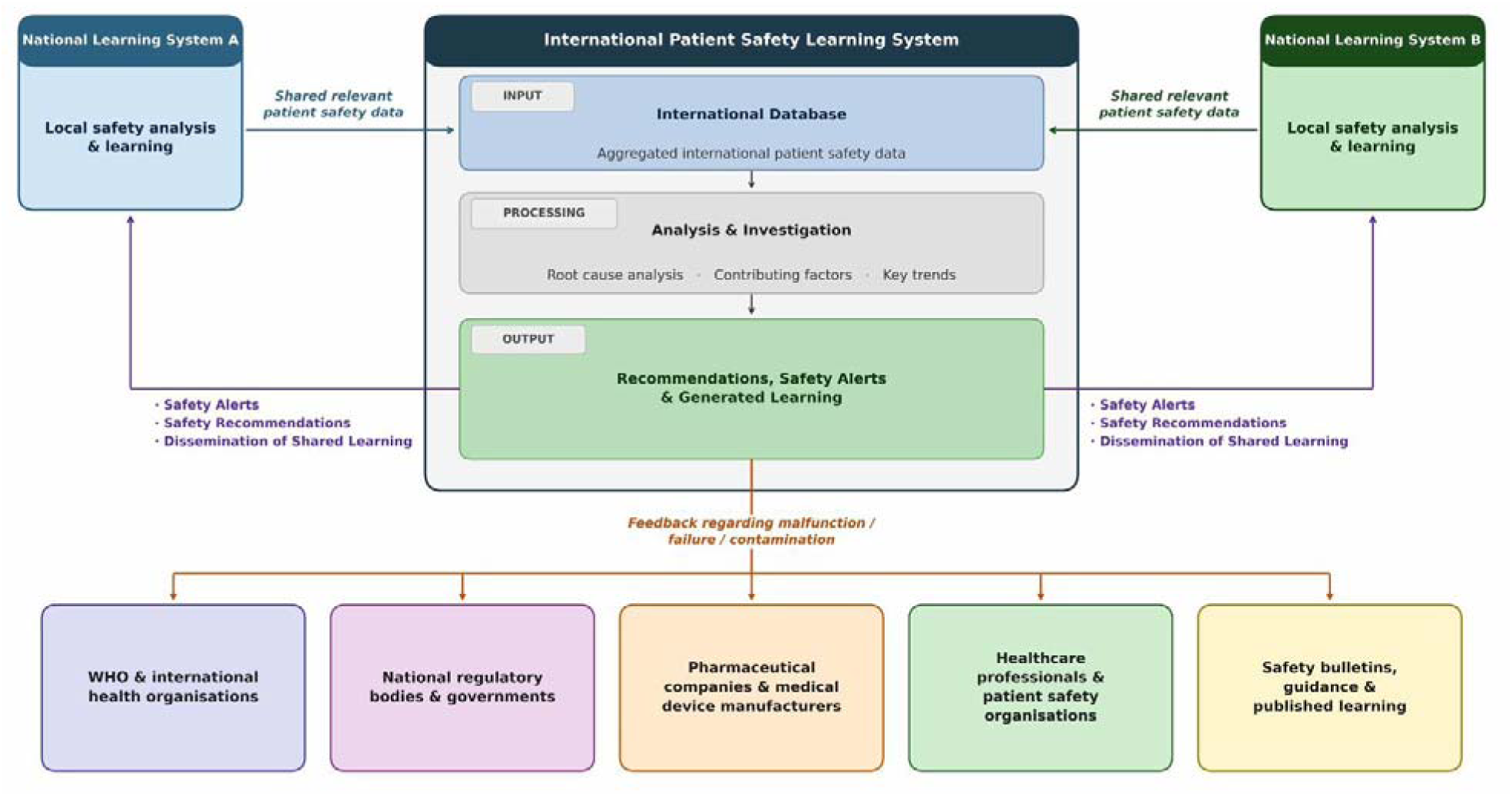
Proposed framework for the design and operationalisation of an international patient safety learning system, based on the combined findings of the systematic literature review, expert interviews, and Delphi consensus. *Note: this figure is best viewed in landscape orientation*.

## DISCUSSION

### Principal findings

This study is the first to establish international healthcare expert consensus on the purposes, key functions and features, and feasibility of a potential international PSLS. The panel reached consensus on 85 of 103 statements across the four domains, endorsing a system designed to facilitate cross-national learning, generate transferable safety recommendations, and coordinate international efforts to mitigate patient safety risks. Importantly, the study produced novel consensus-based lists of patient safety incidents deemed relevant for international sharing and criteria for determining what constitutes a risk of international concern - outputs that have no precedent in the existing literature.

### Comparison with existing literature

This study builds upon the work of Howell et al.,^[34]^ who established consensus on the purposes of national patient safety incident reporting systems. While that study focused on national and local systems, the present study extends this work to the international level and broadens the scope from incident reporting to learning systems more generally. The distinction is important: a learning system can draw on diverse patient safety data sources - including audits, case studies, and investigation reports - rather than relying solely on voluntarily reported incidents, which are subject to well-documented under-reporting.^[43]^

The findings align with the broader literature on international safety learning in other safety-critical industries. In aviation, standardisation of reporting practices and international oversight have been critical to the development of international safety systems.^[42]^ The International Civil Aviation Organization (ICAO) provides a model of how international governance, standardised taxonomy, and mandatory reporting can coexist with mechanisms for learning and feedback across member states. However, the degree to which healthcare can emulate this standardisation remains uncertain, as highlighted by the strong consensus in the present study on the challenge posed by countries being at very different maturity levels with respect to safety culture, and by the established heterogeneity in how national systems collect and classify patient safety data.^[7,8,10]^

The value and limitations of existing national learning systems are illustrated by the case of vincristine-related patient safety incidents in England and Wales. Analysis of 38 reports from the National Reporting and Learning System (NRLS) found that many lacked sufficient narrative detail to characterise the nature of the hazard fully, and that the low volume of such reports - scattered across a large database - risked escaping the attention of regulators and safety experts.^[47]^ Furthermore, deaths following inappropriate vincristine administration persisted over four decades despite repeated guidance, demonstrating how systematic failures - including manufacturing errors, ambiguous labelling, and lack of standardisation - require sustained international coordination rather than local fixes alone.^[46,47]^ This example underscores a key finding of the present study: the importance of generating comprehensive narratives from safety reports, and the value of aggregating data across national systems to surface rare but catastrophic events that no single national system would identify at sufficient volume.

Automation was identified in the present study as both a desirable feature and a source of ongoing challenge. In safety-critical industries more broadly, automation of reporting processes is widespread and continually evolving, but its introduction is rarely straightforward.^[49]^ In healthcare specifically, machine learning approaches applied to patient safety incident reports - particularly those containing large amounts of free text - have shown promise for automated classification of incident content and severity, but substantial development is still required before such tools can reliably support an international system.^[48]^ Physicians’ acceptance of automated systems also cannot be assumed: distrust of automation in clinical contexts has been reported, often arising from insufficient training or misinterpretation of system outputs rather than fundamental opposition to the technology.^[50]^ These considerations are directly relevant to the design of an international PSLS, which must balance the benefits of automation - including consistency and efficiency in data processing - against the risks of generating learning that frontline professionals may not trust or act upon.

### Enablers and barriers to implementation

The enablers identified in the present study are broadly consistent with those required for any large-scale, multi-stakeholder international system. The critical importance of a standardised taxonomy is well established in the literature: without common definitions, aggregation of data across national systems is prone to misclassification and incompatibility.^[7,9]^ The International Classification for Patient Safety (ICPS), developed by the WHO, represents a meaningful attempt to address this, though its adoption across national systems remains incomplete.^[9]^ The panel’s strong endorsement of buy-in from healthcare systems within individual countries and of political will at an international level reflects the established understanding that system-level safety improvements require leadership commitment at multiple levels.^[42]^

Among the barriers, the reporting culture challenge - the degree to which frontline professionals actually use reporting systems - warrants particular attention. Anonymity in reporting processes has been suggested as one mechanism to improve system use, as fear of blame and professional consequences may preclude complete adherence.^[43]^ A tension exists at the frontline between resolving a safety problem locally and reporting it upward: when physicians perceive a safety incident as having been fixed, the impetus to report it formally diminishes, reducing opportunities for system-wide learning.^[51]^ Cultivating a culture in which reporting is habitual - even where local solutions have been identified - is considered a feature of highly reliable healthcare organisations,^[52]^ and this cultural prerequisite must be addressed in any strategy to introduce an international system that depends on consistent national-level reporting as its data input.

The resource implications of establishing and maintaining an international PSLS were acknowledged as a major challenge, with all panellists reaching consensus on funding difficulty and potential costs. The experience of existing international safety learning initiatives suggests that sustainable funding models typically require committed multilateral support, often anchored by an international organisation with an established mandate.^[5,42]^ The WHO, given its role in the Global Patient Safety Action Plan 2021–2030,^[5]^ would be a natural candidate to provide the international governance framework identified by the panel as essential.

### Strengths and limitations

Key strengths of this study include the heterogeneity and seniority of the expert panel, with 21 participants from all six continents representing a combined 314 years of experience in patient safety. The use of successive rounds with structured feedback,^[16,25]^ rigorous predefined consensus criteria, pilot testing, and independent expert validation of the survey instrument contribute to the methodological rigour of the study.

Limitations should be acknowledged. The initial response rate of 29% is lower than some comparable studies, the 21 participants exceeded the predefined minimum panel size of 20, and the 71% retention rate in round two demonstrates strong engagement among those who participated. However, the panel could have benefited from greater representation from non-English-speaking countries and low-resource settings, where an international PSLS may face additional contextual challenges. The post-hoc raising of the consensus threshold from 70% to 80% - while increasing the practical utility of the findings - was not pre-specified and is acknowledged as a limitation. The large number of statements achieving consensus also raises the possibility that the selected statements were not sufficiently contested to require the Delphi method; however, the generation of 41 new statements from qualitative feedback, including the novel subsection on criteria for international concern, demonstrates that the process generated substantive new knowledge. Finally, consensus among experts does not guarantee real-world applicability: the perspectives of frontline end-users are an essential complement to this expert-derived framework.^[17]^

### Implications and future research

The consensus findings provide a foundation for the next phase of research: engaging potential end-users - frontline physicians and other healthcare professionals - to assess the acceptability and usability of the proposed international PSLS. This is essential given that the present panel comprised primarily policymakers, academics, and safety managers rather than frontline clinicians. The proposed framework (figure 3) and the consensus-derived criteria for international sharing offer practical starting points for organisations such as the WHO.^[5]^ The standardisation challenges identified by the panel underscore the need for international collaboration on patient safety taxonomies and data governance frameworks before any implementation can proceed.^[7–9]^ The novel lists of patient safety incidents and criteria for international concern may inform national agencies and international organisations in prioritising which patient safety data should be systematically shared across borders. Future research should also address the governance structures required to operate an international PSLS, including the role of existing international bodies, funding models, and mechanisms for ensuring equitable participation from low- and middle-income countries.

## CONCLUSION

This modified online Delphi study achieved international healthcare expert consensus on the purposes, key functions and features, and feasibility of a potential international patient safety learning system. The findings include novel consensus-based lists of patient safety incidents relevant to international sharing and criteria for determining international concern. The proposed framework provides a platform for future feasibility research and potential pilot implementation, in alignment with the WHO Global Patient Safety Action Plan 2021–2030.^[5]^ Crucially, the findings highlight that structural prerequisites - including a standardised international taxonomy, robust data governance, and equitable participation - must be addressed before any implementation can proceed.

## Supporting information

CREDES Checklist

## DECLARATIONS

### Contributors

JQ designed the study, collected and analysed the data, and drafted the manuscript. ACS conceived the study and supervised all stages. AE and FW contributed to the study design, interpretation of findings, and critical revision of the manuscript. All authors approved the final version.

### Ethics approval

Cardiff University School of Medicine Research Ethics Committee (SMREC Reference Number: 18/70). Consent to participate was obtained electronically from all participants.

### Data availability statement

Anonymised data are available upon reasonable request from the corresponding author.

### Patient and public involvement

A patient representative participated as a member of the expert panel.

### Provenance and peer review

Not commissioned; externally peer reviewed.

## ONLINE SUPPLEMENTARY MATERIALS

**Table S1.** Expert panel inclusion criteria.

**Table S2.** Full ranking of all 103 Delphi statements across both rounds, including percentage agreement, IQR, and group median.

**Table S3.** Newly generated statements from round one qualitative feedback, organised by thematic section.

**Appendix 1.** Delphi survey instrument (rounds one and two).

