## Appendix 1 for "International healthcare experts’ consensus on the key requirements of a potential international patient safety learning system: a modified online Delphi study"

**Delphi Survey Instrument**

*Rounds One and Two*

Qasem J, Edwards A, Wood F, Carson-Stevens A

*Division of Population Medicine, Cardiff University School of Medicine, Cardiff, Wales, United Kingdom*

**Study title:** International healthcare experts’ consensus on the key requirements of a potential international patient safety learning system: a modified online Delphi study

**Ethics approval:** Cardiff University School of Medicine Research Ethics Committee (SMREC Reference Number: 18/70)

**Survey platform:** Qualtrics (Qualtrics, Provo, UT, USA)

**Data collection period:** June–November 2020

**KEY DEFINITIONS**

The following definitions were provided to panellists as hover-over text within the Qualtrics survey platform, accessible by hovering over underlined key terms within each statement. Definitions are reproduced here for transparency.

**International Patient Safety Learning System (PSLS):** A system designed to aggregate, analyse, and share patient safety data and learning across national boundaries, with the aim of identifying internationally relevant patient safety risks and generating transferable safety recommendations.

**Patient safety incident:** Any unintended or unexpected incident that could have or did lead to harm for one or more patients receiving healthcare (World Health Organization, 2009).

**Adverse event:** An incident that results in harm to a patient (World Health Organization, 2009).

**Near miss:** An incident where no harm was done to a patient, but there could have been if people had done the same thing and, if allowed to go a stage further, it could have led to serious harm or death (World Health Organization, 2009).

**Ten times medication error:** A medication dosage error in which a patient receives ten times the prescribed dose of a medication, which can result in serious or fatal harm.

**In vitro diagnostic device (IVD):** A medical device or reagent used to examine specimens derived from the human body, such as blood or tissue, to provide information for diagnosis, monitoring, or prevention of disease.

**Patient Safety Learning System (PSLS):** A system used to collect, analyse, and disseminate information about patient safety incidents and risks to improve safety in healthcare. A learning system may incorporate incident reporting but can also draw on other data sources such as safety audits, case studies, and investigation reports.

**International concern:** A patient safety risk or incident is of international concern when it has the potential to affect patient safety in more than one country and would benefit from coordinated international action, surveillance, or learning.

**ROUND ONE SURVEY**

***Exploring the purpose and feasibility of an international patient safety learning system: A Delphi Study***

*The purpose of this international Delphi study is to achieve expert consensus about the possible expectations from, and the related requirements of an international patient safety learning system.*

*We are seeking your opinion, as part of an anonymous expert panel, to participate in up to three rounds of an online Delphi survey.*

*There are four main sections representing what the literature and key informant interviews outlined to be essential to a potential international patient safety learning system.*

*The sections are as follows:*

- *Section 1: Purpose(s) of an international patient safety learning system*
- *Section 2: Key functions/features of an international patient safety learning system*
- *Section 3: Patient Safety incidents relevant for international sharing and learning*
- *Section 4: Enablers and challenges to the set-up of an international patient safety learning system*

*Note: please refer to Participant Information Sheet for further information, if you have not read it.*

**EXPERIENCE**

**Please note: this section will not identify you personally, and it aims to demonstrate how your experience and expertise contributed to the results of the study.**

| i. | Experience |
| --- | --- |
| i.a | Profession: |
| i.b | Country: |
| i.c | How many years of experience do you have in the field of patient safety/quality improvement (including academic/clinical/other): |
| i.d | Do you have any experience in patient safety at a national/international level? Please give details: |
| i.e | Do you have any experience in leadership roles within healthcare or patient safety organisations? Please give details: |

| ii. | Do you wish to receive feedback regarding the result of this study? | YES (click) | NO (click) |
| --- | --- | --- | --- |

**SECTION 1: PURPOSES OF A POTENTIAL INTERNATIONAL PATIENT SAFETY LEARNING SYSTEM (PSLS)**

**Rating scale (shown once for this section):** Please rate the extent to which you agree that each of the following could be considered a purpose of an international PSLS:

| **1** | **2** | **3** | **4** | **5** | **6** | **7** | **8** | **9** |
| --- | --- | --- | --- | --- | --- | --- | --- | --- |
| *Strongly disagree* |  |  |  | *Neutral* |  |  |  | *Strongly agree* |

*Ratings of 1–3 = Disagree (this is not a purpose of an international PSLS); 4–6 = Neutral; 7–9 = Agree (this is a purpose of an international PSLS).*

**1.01** Identification of patient safety risks relevant internationally.

**1.02** Surveillance of patient safety incidents to detect potential risks relevant internationally.

**1.03** Process learning from investigations of patient safety incidents so that transferable learning between countries can be identified.

**1.04** Generate systems improvement strategies based on documented efforts to mitigate risk in other countries.

**1.05** Learn from reported common patient safety risks to coordinate efforts internationally to mitigate and address those risks.

**1.06** Learn from reported common patient safety risks internationally and work with other countries to innovate solutions to prevent those risks from reaching patients in similar healthcare contexts.

**1.07** Awareness of frequently occurring patient safety incidents in other countries.

**1.08** Drive up standards in learning from patient safety incidents.

**1.09** Standardise the way learning from investigating patient safety incidents is reported.

**1.10** Coordinate the design of initiatives/interventions to mitigate commonly identified patient safety risk.

**1.11** Provide an overview of the most important risk areas and be able to learn with and from other countries.

**1.12** Focus on assimilating learning about patient safety incidents that provide serious and specific insights into system safety.

*Where you have selected a value of 6 or below, please justify your answer(s).*

*Optional: Are there any changes or additions to the stated purposes that you think should be included? Please provide your suggestion(s).*

**SECTION 2: KEY FEATURES AND FUNCTIONS OF A POTENTIAL INTERNATIONAL PSLS**

**Rating scale:** Please rate the extent to which you agree that each of the following should be a key feature or function of an international PSLS:

| **1** | **2** | **3** | **4** | **5** | **6** | **7** | **8** | **9** |
| --- | --- | --- | --- | --- | --- | --- | --- | --- |
| *Strongly disagree* |  |  |  | *Neutral* |  |  |  | *Strongly agree* |

*Ratings of 1–3 = Disagree (this is not a key feature/function); 4–6 = Neutral; 7–9 = Agree (this is a key feature/function).*

**2.01** Generate safety recommendations with solutions that can be shared with countries and considered for adoption in different contexts in healthcare systems.

**2.02** Collate patient safety data/reports/resources in the interest of enabling international-level learning.

**2.03** Coordinate and launch international-level efforts to tackle common patient safety risks.

**2.04** A proactive approach for identifying patient safety risks that require international action.

**2.05** Ability to analyse and make recommendations based on shared patient safety data to identify transferable learning from countries responding to patient safety risks.

**2.06** Ability to use filters to search multiple sources of data/evidence/outputs to get to required information quickly.

**2.07** A repository for reports describing how interventions to mitigate risk work and how to support others to support implementation elsewhere.

**2.08** Ability to trigger an alarm that deploys a notification to countries about serious identified patient safety risks.

**2.09** Compile and consolidate information from multiple countries.

**2.10** Analyse information and generate reports based on inputs from multiple countries and consolidating the ‘meta-learning’ (i.e., the overall learning from the country-level learning).

**2.11** Ability to share reported patient safety incidents/risks relevant to international learning.

**2.12** Shared database of resources that could support risk mitigation/prevention.

**2.13** Support improvements in collection of patient safety data through sharing exemplars.

**2.14** Support improvement in incident reporting and learning through sharing exemplars.

**2.15** Develop instructional manuals on how to investigate and learn from patient safety incidents.

**2.16** In the case of patient safety incidents related to equipment and/or medication, manufacturers will be informed to take action to mitigate and contain any further risk.

**2.17** Ability to identify contributing factors to patient safety risks that can be transversal to certain types of incidents, contexts, and case studies.

*Where you have selected a value of 6 or below, please justify your answer(s).*

*Optional: Are there any changes or additions to the stated features and functions that you think should be included? Please provide your suggestion(s).*

**SECTION 3: PATIENT SAFETY INCIDENTS RELEVANT TO INTERNATIONAL SHARING AND LEARNING**

*There have been examples where researchers from multiple countries have pooled patient safety incident report data to maximise opportunities to learn from unsafe care. If a potential international mechanism existed which permitted confidential, information-governance compliant data sharing, which types of patient safety incidents could be relevant and essential for international learning.*

**Rating scale:** Please rate the extent to which you agree that each of the following types of patient safety incident should be relevant to an international PSLS for the purposes of international sharing and learning:

| **1** | **2** | **3** | **4** | **5** | **6** | **7** | **8** | **9** |
| --- | --- | --- | --- | --- | --- | --- | --- | --- |
| *Strongly disagree* |  |  |  | *Neutral* |  |  |  | *Strongly agree* |

*Ratings of 1–3 = Disagree (this incident type is not relevant internationally); 4–6 = Neutral; 7–9 = Agree (this incident type is relevant internationally).*

**3.01** All adverse events (an incident that results in harm to a patient) (WHO, 2009).

**3.02** A near miss (events where no harm was done, but there could have been if people had done the same thing and, if allowed to go a stage further, it could have led to a death) (WHO, 2009).

**3.03** Incidents relevant to current international campaigns or challenges (e.g., the WHO patient safety challenges).

**3.04** Incidents identified by multiple countries as common sources of unsafe care and therefore a potential priority to learn from and tackle collectively.

**3.05** Tubing misconnections/misconnection errors (e.g., non-luer connected devices).

**3.06** Nasogastric tube positioning errors/incidents.

**3.07** Ten times medication errors (refer to definition provided above).

**3.08** Incidents related to manufacturing and supply chain (e.g., contaminated vaccine/IV fluids/injections/drugs).

**3.09** Incidents related to faulty medical devices (including IVDs) or equipment failure.

*Where you have selected a value of 6 or below, please justify your answer(s).*

*Open question: Are there any changes or additions to the stated patient safety incidents that you would like to make/add? If so, please provide your own changes.*

*In your opinion, what are the criteria for deciding whether an incident is an international concern (global priority), particularly for international learning?*

**SECTION 4: ENABLERS AND CHALLENGES TO SETTING UP AN INTERNATIONAL PATIENT SAFETY LEARNING SYSTEM (PSLS)**

*When planning to set up a learning system, especially at the international level, there are potential challenges and enabling factors that could support or hinder the process of setting up such as system. Anticipating some of these factors beforehand would help in determining the feasibility of such a system in the future.*

**SECTION 4A: KEY ENABLERS TO SETTING UP AN INTERNATIONAL PSLS**

**Rating scale:** Please rate the extent to which you agree that each of the following would be a key enabler to setting up an international PSLS:

| **1** | **2** | **3** | **4** | **5** | **6** | **7** | **8** | **9** |
| --- | --- | --- | --- | --- | --- | --- | --- | --- |
| *Strongly disagree* |  |  |  | *Neutral* |  |  |  | *Strongly agree* |

*Ratings of 1–3 = Disagree (this is not a key enabler); 4–6 = Neutral; 7–9 = Agree (this is a key enabler).*

**4.01** Governments, agencies, and organisations have clear legislation, regulations and guidelines that are supportive of and encouraging of sharing of patient safety data relevant for international learning.

**4.02** An independent body/organisation responsible for operating and maintaining the international PSLS.

**4.03** Country-level will to learn with and from other countries.

**4.04** Funded by partner countries.

**4.05** Having buy-in from healthcare systems within each individual country.

**4.06** Having buy-in from international organisations with a role and interest in patient safety.

**4.07** Broad support from the clinicians and the societies that make up the international patient safety community.

**4.08** Involvement of important knowledge mobilisers (e.g., Institute for Healthcare Improvement, World Health Organization).

**4.09** To have representation from countries that are submitting data to the international system (e.g., national co-ordinators).

**4.10** Existence of international standards/convention for sharing patient safety data.

**4.11** For users and/or contributors to the system to be able to access it in an easy, secure way.

**4.12** Multiple input methods are available (e.g., online, phone, e-mail, etc.).

**4.13** The system is accessible for everyone to read the information that helps organisations and individuals learn (e.g., web page open for everyone).

**4.14** Access for input to the system should be limited and discussed nationally, with only one named responsible organisation in each country.

**4.15** Password-controlled access to an online platform.

**4.16** Automation of as many functions as possible.

*Where you have selected a value of 6 or below, please justify your answer(s).*

*Optional: Are there any changes or additions to the stated enabling factors that you would like to make? If so, please provide your own changes.*

**SECTION 4B: KEY CHALLENGES AND BARRIERS TO SETTING UP AN INTERNATIONAL PSLS**

**Rating scale:** Please rate the extent to which you agree that each of the following would be a key challenge or barrier to setting up an international PSLS:

| **1** | **2** | **3** | **4** | **5** | **6** | **7** | **8** | **9** |
| --- | --- | --- | --- | --- | --- | --- | --- | --- |
| *Strongly disagree* |  |  |  | *Neutral* |  |  |  | *Strongly agree* |

*Ratings of 1–3 = Disagree (this is not a key barrier); 4–6 = Neutral; 7–9 = Agree (this is a key barrier).*

**4.17** Finding the responsible international organisation to set up the system and relevant national organisations that will commit to co-operate.

**4.18** At the international level, big cultural factors and different types of healthcare systems might present a challenge.

**4.19** Lack of a sense of ownership in a system the end-user has not been involved in developing.

**4.20** Having too many different bodies which are involved.

**4.21** Limited resources in less developed countries.

**4.22** Issues about national reputation, healthcare system reputation issues, which might create barriers.

**4.23** Concerns regarding privacy issues when sharing learning from investigations of patient safety incidents.

**4.24** Lack of common format/approach to sharing learning about patient safety risks and/or efforts to mitigate/prevent harm to patients.

*Where you have selected a value of 6 or below, please justify your answer(s).*

*Optional: Are there any changes or additions to the stated challenges/barriers that you would like to make? If so, please provide your own changes.*

**ROUND TWO SURVEY**

***Exploring the purpose and feasibility of an international patient safety learning system: A Delphi Study***

*In this second round of the Delphi study you will have the opportunity to revise your rating for statements from the first round, and you will also be able to rate new statements suggested by fellow panellists, including yourself.*

*The sections for this round are as follows:*

- *Section 1: Purpose(s) of an international patient safety learning system*
- *Section 2: Key functions/features of an international patient safety learning system*
- *Section 3: A - Patient Safety incidents relevant to international sharing and learning*

*B – Criteria for deciding what patient safety risk is of international concern*

- *Section 4: Enablers and challenges to the set-up of an international patient safety learning system*

**SECTION 1: PURPOSES OF A POTENTIAL INTERNATIONAL PSLS**

**Rating scale:** Please rate the extent to which you agree that each of the following could be a purpose of an international PSLS:

| **1** | **2** | **3** | **4** | **5** | **6** | **7** | **8** | **9** |
| --- | --- | --- | --- | --- | --- | --- | --- | --- |
| *Strongly disagree* |  |  |  | *Neutral* |  |  |  | *Strongly agree* |

*Statements marked with ▶ are carried forward from round one (not yet at consensus). Statements marked with ★ are new, generated from round one feedback.*

**1.01 ▶** Awareness of frequently occurring patient safety incidents in other countries.

**1.02 ▶** Standardise the way learning from investigating patient safety incidents is reported.

**1.03 ▶** Coordinate the design of initiatives/interventions to mitigate commonly identified patient safety risk.

**1.04 ★** Identify evidence-practice gaps that are occurring in similar contexts in other countries in order to develop interventions together.

**1.05 ★** Learn from patients and families about risk, harm, response, and remediation.

**1.06 ★** Bolster capability of those seeking to improve safety by benefitting from the shared learnings of those that have achieved improvements in safety in similar care contexts.

**1.07 ★** Learn with and from countries about efforts to improve patient safety in terms of what works and how.

**1.08 ★** Identify priorities for research and development to focus international efforts and resources where they are needed most.

**1.09 ★** Co-ordinate efforts of national patient safety organisations in collaboration with the WHO.

**1.10 ★** Sharing affordable design-based interventions to reduce patient safety risks.

*Where you have selected a value of 6 or below, please justify your answer(s).*

*Optional: Are there any changes or additions to the stated purposes that you think should be included? Please provide your suggestion(s).*

**SECTION 2: KEY FEATURES AND FUNCTIONS OF A POTENTIAL INTERNATIONAL PSLS**

**Rating scale:** Please rate the extent to which you agree that each of the following should be a key feature or function of an international PSLS:

| **1** | **2** | **3** | **4** | **5** | **6** | **7** | **8** | **9** |
| --- | --- | --- | --- | --- | --- | --- | --- | --- |
| *Strongly disagree* |  |  |  | *Neutral* |  |  |  | *Strongly agree* |

*Statements marked with ▶ are carried forward from round one. Statements marked with ★ are new.*

**2.01 ▶** Ability to use filters to search multiple sources of data/evidence/outputs to get to required information quickly.

**2.02 ▶** Develop instructional manuals on how to investigate and learn from patient safety incidents.

**2.03 ▶** Ability to identify contributing factors to patient safety risks that can be transversal to certain types of incidents, contexts, and case studies.

**2.04 ★** Facilitate learning and support through a range of formats (e.g., instructional manuals, train-the-trainer workshops, webinars, onsite learning).

**2.05 ★** Develop a structured process for investigating patient safety risks identified by the international learning system.

**2.06 ★** Develop methodologies to evaluate practices to learn from patients and family members in all facets of patient safety, including prevention, response, recovery, and remediation.

**2.07 ★** Support the progression of countries towards a proactive and personalised clinical risk management approach (i.e., moving from a reactive to a proactive mindset).

**2.08 ★** To develop methodological approaches for assimilating a range of descriptive patient safety data to build a more complete understanding of safety within countries.

*Where you have selected a value of 6 or below, please justify your answer(s).*

*Optional: Are there any changes or additions to the stated features and functions that you think should be included? Please provide your suggestion(s).*

**SECTION 3: PATIENT SAFETY INCIDENTS RELEVANT TO INTERNATIONAL SHARING AND LEARNING**

*Knowing types of patient safety incidents/risks that are essential for international sharing and learning is helpful. Therefore, knowing the criteria for deciding what patient safety risk is of international concern is key to gain maximum benefit from the international PSLS.*

**SECTION 3A: PATIENT SAFETY INCIDENTS RELEVANT TO INTERNATIONAL SHARING**

**Rating scale:** Please rate the extent to which you agree that each of the following types of patient safety incident should be relevant to an international PSLS for the purposes of international sharing and learning:

| **1** | **2** | **3** | **4** | **5** | **6** | **7** | **8** | **9** |
| --- | --- | --- | --- | --- | --- | --- | --- | --- |
| *Strongly disagree* |  |  |  | *Neutral* |  |  |  | *Strongly agree* |

*Statements marked with ▶ are carried forward from round one. Statements marked with ★ are new.*

**3.01 ▶** All adverse events (an incident that results in harm to a patient) (WHO, 2009).

**3.02 ▶** A near miss (events where no harm was done, but there could have been if people had done the same thing and, if allowed to go a stage further, it could have led to a death) (WHO, 2009).

**3.03 ★** Incidents where risk of severe harm or death is likely, should the same incident reoccur.

**3.04 ★** Incidents that result from relatively novel contributory factors (e.g., pandemics, electricity outage, internet outage, civil unrest, war, funding issues, natural disasters, deliberate sabotage, criminal activity).

**3.05 ★** Patient safety risks that are new and/or have not been reported before in your country.

**3.06 ★** Patient safety risks related to diagnostic errors.

**3.07 ★** Incidents related to drug and equipment safety.

**3.08 ★** Any incident type that impacts paediatric patients.

**3.09 ★** Incident involves a product or device that might be a major contributing factor to the incident.

*Where you have selected a value of 6 or below, please justify your answer(s).*

*Optional: Are there any changes or additions to the stated patient safety incidents that you would like to make/add? If so, please provide your own changes.*

**SECTION 3B: CRITERIA FOR DECIDING WHAT PATIENT SAFETY RISK IS OF INTERNATIONAL CONCERN**

**Rating scale:** Please rate the extent to which you agree that each of the following should be a criterion for determining whether a patient safety risk is of international concern:

| **1** | **2** | **3** | **4** | **5** | **6** | **7** | **8** | **9** |
| --- | --- | --- | --- | --- | --- | --- | --- | --- |
| *Strongly disagree* |  |  |  | *Neutral* |  |  |  | *Strongly agree* |

**3.10** Risk of harming a large number of individuals in multiple countries if no intervention is taken.

**3.11** Morbidity and mortality from patient safety risks/events (i.e., impact and severity).

**3.12** The proposed patient safety solution needs international action (e.g., action from major pharmaceutical companies).

**3.13** Identified patient safety risk(s) is/are relevant to more than one country facing similar clinical challenges (e.g., prevention and control of infectious disease).

**3.14** Identified patient safety risk(s) is/are relevant to more than one country (e.g., supply of material/medicine/raw materials/devices originating in another country).

**3.15** Ease of measurement of the identified patient safety risk(s) in multiple countries.

**3.16** Stakeholders’ interest in specific patient safety incidents/risks.

**3.17** The availability of evidence to support unequivocal preventability.

*Where you have selected a value of 6 or below, please justify your answer(s).*

*Optional: Are there any changes or additions to the stated patient safety incident criteria that you would like to make/add? If so, please provide your own changes.*

**SECTION 4: ENABLERS AND CHALLENGES TO SETTING UP AN INTERNATIONAL PATIENT SAFETY LEARNING SYSTEM (PSLS)**

*When planning to set up a learning system, especially at the international level, there are potential challenges and enabling factors that could support or hinder the process of setting up such as system. Anticipating some of these factors beforehand would help in determining the feasibility of such a system in the future.*

**SECTION 4A: KEY ENABLERS TO SETTING UP AN INTERNATIONAL PSLS**

**Rating scale:** Please rate the extent to which you agree that each of the following would be a key enabler to setting up an international PSLS:

| **1** | **2** | **3** | **4** | **5** | **6** | **7** | **8** | **9** |
| --- | --- | --- | --- | --- | --- | --- | --- | --- |
| *Strongly disagree* |  |  |  | *Neutral* |  |  |  | *Strongly agree* |

*Statements marked with ▶ are carried forward from round one. Statements marked with ★ are new.*

**4.01 ▶** Access for input to the system should be limited and discussed nationally, with only one named responsible organisation in each country.

**4.02 ▶** Password-controlled access to an online platform.

**4.03 ★** Having political will to generate and co-ordinate patient safety interventions of international concern.

**4.04 ★** Having internationally comparative patient safety data.

**4.05 ★** Deploying resources/learning where they are needed most.

**4.06 ★** Encouraging and funding cross-jurisdictional studies of strategies designed and evaluated to understand better system functioning and implementation impact.

**4.07 ★** The benefits for those who are supposed to feed the information to the system are very clear.

**4.08 ★** Having the patient experience as a facilitator around which common definitions of harm and their priority for mitigation are set.

**4.09 ★** Having a standardised international patient safety taxonomy.

*Where you have selected a value of 6 or below, please justify your answer(s).*

*Optional: Are there any changes or additions to the stated enabling factors that you would like to make? If so, please provide your own changes.*

**SECTION 4B: KEY CHALLENGES AND BARRIERS TO SETTING UP AN INTERNATIONAL PSLS**

**Rating scale:** Please rate the extent to which you agree that each of the following would be a key challenge or barrier to setting up an international PSLS:

| **1** | **2** | **3** | **4** | **5** | **6** | **7** | **8** | **9** |
| --- | --- | --- | --- | --- | --- | --- | --- | --- |
| *Strongly disagree* |  |  |  | *Neutral* |  |  |  | *Strongly agree* |

**4.10** Difficulty with funding this learning system even from participating countries.

**4.11** Potential cost of establishing and maintaining the system.

**4.12** Lack of a data/information governance strategy.

**4.13** The “what’s in it for me” problem, i.e., to find the value proposition/business case in the various and varied jurisdictions.

**4.14** Many countries have multiple reporting and learning organisations.

**4.15** Many countries are at very different maturity levels with respect to just/safety culture.

**4.16** The information collected by this system will have limited utility if it is relying on system-centric versions of patient harm without explicitly including the patient perspective of harm.

*Where you have selected a value of 6 or below, please justify your answer(s).*

*Optional: Are there any changes or additions to the stated challenges/barriers that you would like to make? If so, please provide your own changes.*
