## Supplementary material for "International healthcare experts’ consensus on the key requirements of a potential international patient safety learning system: a modified online Delphi study": CREDES Checklist

**Guidance on Conducting and REporting DElphi Studies**

*Jünger S, Payne SA, Brine J, Radbruch L, Brearley SG. Palliative Medicine 2017;31(8):684–706.*

| ☑ | Item is reported in the manuscript |
| --- | --- |
| ☐ | Item is not applicable or not reported (see Location column for explanation) |

| **Domain** | **Item** | **Requirement** | **☑** | **Location in manuscript** |
| --- | --- | --- | --- | --- |
| **1. Rationale for the Delphi Technique** | | | | |
| **Justification** | **Rationale for method** | The choice of the Delphi technique as a method of systematically collating expert consultation and building consensus needs to be well justified. | ☑ | *Introduction, final paragraph; Methods — Study design. Rationale given: need to validate SLR and interview findings with healthcare experts, international scope, geographically dispersed panel.* |
| **2. Planning and Design** | | | | |
| **Planning and process** | **Modifications justified** | The Delphi technique is a flexible method and can be adjusted to the respective research aims and purposes. Any modifications should be justified by a rationale and be applied systematically and rigorously. | ☑ | *Methods — Study design. Modification: systematic literature review and semi-structured interviews replaced the traditional first qualitative round; justification provided. Described as a ‘modified online Delphi’.* |
| **Definition of consensus** | **A priori consensus criteria defined** | An a priori criterion for consensus should be defined, including: (a) how to proceed with items in the next round; (b) the threshold to terminate the Delphi; and (c) procedures when consensus is (not) reached. | ☑ | *Methods — Consensus definition and termination criteria. Defined a priori: ≥70% agreement (ratings 7–9) with IQR ≤2.00. Termination if ≥80% of all statements reached consensus or round 2 response rate <70%. Post-hoc raising to ≥80% threshold acknowledged as a limitation.* |
| **3. Study Conduct** | | | | |
| **Informational input** | **Materials piloted and reviewed** | All material provided to the expert panel at the outset and throughout the Delphi process should be carefully reviewed and piloted in advance to examine the effect on experts’ judgements and to prevent bias. | ☑ | *Methods — Survey development. Survey piloted with two subject matter experts; validated by WHO Collaborating Centre experts in Florence, Italy. Changes made to layout, wording, consent form, and estimated completion time.* |
| **Prevention of bias** | **Measures to avoid influencing experts** | Researchers need to take measures to avoid directly or indirectly influencing the experts’ judgements. If one or more members of the research team have a conflict of interest, an independent researcher should coordinate the study. | ☑ | *Methods — Delphi procedure. Anonymity maintained between panellists throughout; de-identified group results only provided to panellists. Declarations: no competing interests declared by any author.* |
| **Interpretation and processing of results** | **Non-consensus interpreted** | Consensus does not necessarily imply the ‘correct’ answer; (non-)consensus and stable disagreement provide informative insights and highlight differences in perspectives. | ☑ | *Results and Discussion. Non-consensus statements identified and discussed (e.g., automated alerting, access restrictions, funding by partner countries). Post-hoc threshold change and its implications discussed in Limitations.* |
| **External validation** | **Final guidance reviewed externally** | It is recommended to have the final draft of the resulting guidance reviewed and approved by an external board or authority before publication. | ☑ | *Methods — Survey development. Survey instrument reviewed and validated by experts at the WHO Collaborating Centre in Human Factors and Communication, Florence, Italy prior to round one. Supervisory team provided ongoing critical oversight throughout.* |
| **4. Reporting** | | | | |
| **Purpose and rationale** | **Purpose clearly defined** | The purpose of the study should be clearly defined and demonstrate the appropriateness of the Delphi technique. A rationale for the choice of the Delphi technique as the most suitable method needs to be provided. | ☑ | *Introduction (final paragraph) and Methods — Study design. Aim and four objectives stated. Delphi selected for anonymous structured consensus-building among geographically dispersed international experts.* |
| **Expert panel** | **Selection criteria and recruitment described** | Criteria for the selection of experts and transparent information on recruitment of the expert panel, sociodemographic details including expertise regarding the topic in question, and (non-)response and response rates over rounds. | ☑ | *Methods — Expert selection and recruitment; Results — Panel characteristics; Table 2 (panel demographics); Online supplementary Table S1 (inclusion criteria); Figure 1 (flowchart). Response rates: 29% round 1, 71% retention round 2.* |
| **Description of methods** | **Methods comprehensively described** | The methods employed need to be comprehensible; this includes preparatory steps, piloting, survey instrument design, number of rounds, data analysis, processing and synthesis of experts’ responses, and methodological decisions taken throughout. | ☑ | *Methods section in full (Study design; Expert selection; Survey development; Delphi procedure; Consensus definition; Data analysis). Appendix 1 provides full survey instrument for both rounds.* |
| **Procedure** | **Flowchart of Delphi stages provided** | A flow chart should illustrate the stages of the Delphi process, including the preparatory phase, actual Delphi rounds, interim steps of data processing and analysis, and concluding steps. | ☑ | *Figure 1. Flowchart illustrates expert identification, screening, invitation, and participation across both rounds including dropout and snowball sampling.* |
| **Definition and attainment of consensus** | **Consensus process comprehensible** | It needs to be comprehensible to the reader how consensus was achieved throughout the process, including strategies to deal with non-consensus. | ☑ | *Methods — Consensus definition and termination criteria; Results — Overview of consensus. Table 1 reports consensus rates by section. Supplementary Table S2 reports per-statement statistics (% agreement, IQR, median) for all 103 statements across both rounds.* |
| **Results** | **Results reported per round** | Reporting of results for each round separately is highly advisable to make the evolving of consensus transparent. This includes average group responses, changes between rounds, and modifications to the survey instrument. | ☑ | *Results section. Round 1 and Round 2 results reported separately within each subsection. Consensus rates per round noted. New statements generated from round 1 qualitative feedback clearly distinguished in Round 2 reporting and in Appendix 1.* |
| **Discussion of limitations** | **Limitations critically reflected** | Reporting should include a critical reflection of potential limitations and their impact on the resulting guidance. | ☑ | *Discussion — Strengths and limitations. Limitations addressed: 29% round 1 response rate; limited representation from non-English-speaking and low-resource settings; post-hoc consensus threshold change; risk of insufficient contestation of statements; expert panel not representative of frontline end-users.* |
| **Adequacy of conclusions** | **Conclusions reflect scope and applicability** | The conclusions should adequately reflect the outcomes of the Delphi study with a view to the scope and applicability of the resulting practice guidance. | ☑ | *Conclusion and Discussion — Implications and future research. Conclusions scoped to expert consensus only; explicitly note need for validation with frontline end-users before implementation; structural prerequisites highlighted.* |
| **Publication and dissemination** | **Guidance identifiable; dissemination planned** | The resulting guidance should be clearly identifiable from the publication, including recommendations for transfer into practice and implementation. Dissemination plan should include endorsement by professional associations and health authorities. | ☑ | *Discussion — Implications and future research. Framework (Figure 3) and consensus-derived criteria presented as starting points for WHO and national agencies. medRxiv pre-print planned prior to peer-reviewed publication to maximise open-access dissemination.* |

**Notes on CREDES applicability:**

The CREDES checklist was developed primarily for Delphi studies used to generate best practice guidance in healthcare, originally within palliative care (Jünger et al., 2017). The present study applies the Delphi technique to achieve expert consensus on the design requirements of a system rather than on clinical practice guidance. All applicable items have been addressed. The item on ‘external validation’ is addressed through WHO Collaborating Centre expert review of the survey instrument; no formal external endorsement of a clinical guideline was applicable given the nature of the study outputs.

The post-hoc raising of the consensus threshold from 70% to 80% is noted as a deviation from the pre-specified protocol and is acknowledged as a limitation in the manuscript.

**Reference:** Jünger S, Payne SA, Brine J, Radbruch L, Brearley SG. Guidance on Conducting and REporting DElphi Studies (CREDES) in palliative care: Recommendations based on a methodological systematic review. Palliative Medicine. 2017;31(8):684–706. doi:10.1177/0269216317690685
