## Supplementary Material for "International healthcare experts’ consensus on the key requirements of a potential international patient safety learning system: a modified online Delphi study"

This document contains four supplementary tables:

- Table S1: Inclusion criteria for the expert panel
- Table S2: Full ranking of all Delphi statements across both rounds (103 statements)
- Table S3: Newly generated statements from round one qualitative feedback, included in round two

**Table S1: Inclusion criteria for the expert panel**

Experts were invited to participate in the first round of the Delphi study if they met the criteria below. Participants identified via snowball sampling were included only where they met all applicable inclusion criteria.

| **Category** | **Inclusion Criteria** |
| --- | --- |
| **Publication-based criteria** | Listed author in a peer-reviewed publication (1999–2020, English language) relevant to: (1) Patient safety; (2) Incident reporting and learning systems; or (3) Organisational learning/knowledge mobilisation. |
| **Experience-based criteria (at least one required)** | Expertise in the development, management, or evaluation of an incident reporting and learning system. |
|  | A role at a local, national, or international level in patient safety and/or incident reporting and learning systems. |
|  | Nominated by a participating panel member (snowball sampling), subject to meeting all other inclusion criteria. |
| **Essential (all participants)** | Active, publicly available e-mail address to enable participation in the online Delphi survey. |

**Table S2: Full ranking of all Delphi statements**

Statements are ranked within each section by percentage agreement (descending), then IQR (ascending), then group median (descending). Statements highlighted in light red did not achieve consensus at the ≥80% agreement threshold (applied post hoc). All statements were rated on a 9-point Likert-like scale (1 = not at all important; 9 = extremely important). Consensus was defined as ≥70% agreement (ratings 7–9) with IQR ≤2.00; a post-hoc threshold of ≥80% agreement was applied to identify the strongest areas of agreement.

| **Ref.** | **Statement** | **% Agreement** | **% Neutral** | **% Disagree-ment** | **IQR** | **Group Median** |
| --- | --- | --- | --- | --- | --- | --- |
| **Section 1: Purpose(s) of a potential international patient safety learning system (PSLS)** | | | | | | |
| 1.01 | Identify priorities for research and development to focus international efforts and resources where they are needed most. | 100% | 0% | 0% | 1.0 | 8 |
| 1.02 | Identify evidence-practice gaps that are occurring in similar contexts in other countries in order to develop interventions together. | 100% | 0% | 0% | 1.0 | 7 |
| 1.03 | Sharing affordable design-based interventions. | 100% | 0% | 0% | 1.5 | 8 |
| 1.04 | Learn with and from countries about efforts to improve patient safety in terms of what works and how. | 93% | 7% | 0% | 1.5 | 8 |
| 1.05 | Process learning from investigations of patient safety incidents so that transferable learning between countries can be identified. | 90% | 10% | 0% | 1.0 | 9 |
| 1.06 | Drive up standards in learning from patient safety incidents. | 90% | 5% | 5% | 1.0 | 9 |
| 1.07 | Identification of patient safety risks relevant internationally. | 90% | 10% | 0% | 2.0 | 8 |
| 1.08 | Awareness of frequently occurring patient safety incidents in other countries. | 87% | 7% | 7% | 2.0 | 8 |
| 1.09 | Generate systems improvement strategies based on documented efforts to mitigate risk in other countries. | 86% | 14% | 0% | 2.0 | 8 |
| 1.10 | Focus on assimilating learning about patient safety incidents that provide serious and specific insights into system safety. | 86% | 14% | 0% | 2.0 | 8 |
| 1.11 | Learn from reported common patient safety risks to coordinate efforts internationally to mitigate and address those risks. | 81% | 14% | 5% | 1.0 | 8 |
| 1.12 | Provide an overview of the most important risk areas and be able to learn with and from other countries. | 81% | 19% | 0% | 1.0 | 8 |
| 1.13 | Learn from reported common patient safety risks internationally and work with other countries to innovate solutions to prevent those risks from reaching patients in similar healthcare contexts. | 81% | 14% | 5% | 2.0 | 8 |
| 1.14 | Co-ordinate efforts of national patient safety organisations in collaboration with WHO. | 80% | 20% | 0% | 1.0 | 8 |
| 1.15 | Bolster capability of those seeking to improve safety by benefitting from the shared learnings of those that have achieved improvements in safety in similar care contexts. | 80% | 20% | 0% | 2.0 | 8 |
| 1.16 | *Surveillance of patient safety incidents to detect potential risks relevant internationally.* | 76% | 19% | 5% | 2.0 | 8 |
| 1.17 | *Learn from patients and families about risk, harm, response, and remediation.* | 73% | 27% | 0% | 1.5 | 8 |
| 1.18 | *Coordinate the design of initiatives/interventions to mitigate commonly identified patient safety risk.* | 73% | 27% | 0% | 1.5 | 7 |
| 1.19 | *Standardise the way learning from investigating patient safety incidents is reported.* | 67% | 27% | 7% | 2.0 | 7 |
| **Section 2: Key features and functions of a potential international PSLS** | | | | | | |
| 2.01 | Develop a structured process for investigating patient safety risks identified by the international learning system. | 100% | 0% | 0% | 2.0 | 7 |
| 2.02 | In the case of patient safety incidents related to equipment and/or medication, manufacturers will be informed to take action to mitigate and contain any further risk. | 95% | 5% | 0% | 1.0 | 9 |
| 2.03 | Generate safety recommendations with solutions that can be shared with countries and considered for adoption in different contexts in healthcare systems. | 95% | 5% | 0% | 2.0 | 8 |
| 2.04 | To develop methodological approaches for assimilating a range of descriptive patient safety data to build a more complete understanding of safety within countries. | 93% | 7% | 0% | 1.0 | 8 |
| 2.05 | Support the progression of countries towards a proactive and personalised clinical risk management approach (i.e. moving from a reactive to a proactive mindset). | 93% | 7% | 0% | 1.5 | 8 |
| 2.06 | Ability to use filters to search multiple sources of data/evidence/outputs to get to required information quickly. | 87% | 7% | 7% | 0.5 | 8 |
| 2.07 | Ability to identify contributing factors to patient safety risks that can be transversal to certain types of incidents, contexts, and case studies. | 87% | 7% | 7% | 1.0 | 8 |
| 2.08 | Facilitate learning and support through a range of formats (e.g. instructional manuals, train-the-trainer workshops, webinars, onsite learning). | 87% | 13% | 0% | 1.0 | 7 |
| 2.09 | Develop methodologies to evaluate practices to learn from patients and family members in all facets of patient safety, including prevention, response, recovery, and remediation. | 87% | 13% | 0% | 1.5 | 7 |
| 2.10 | Collate patient safety data/reports/resources in the interest of enabling international-level learning. | 86% | 14% | 0% | 2.0 | 8 |
| 2.11 | Coordinate and launch international-level efforts to tackle common patient safety risks. | 86% | 14% | 0% | 2.0 | 8 |
| 2.12 | Shared database of resources that could support risk mitigation/prevention. | 86% | 10% | 5% | 2.0 | 8 |
| 2.13 | A proactive approach for identifying patient safety risks that require international action. | 81% | 19% | 0% | 2.0 | 8 |
| 2.14 | Compile and consolidate information from multiple countries. | 81% | 19% | 0% | 2.0 | 8 |
| 2.15 | A repository for reports describing how interventions to mitigate risk work and how to support others to support implementation elsewhere. | 81% | 19% | 0% | 2.0 | 8 |
| 2.16 | Analyse information and generate reports based on inputs from multiple countries and consolidating the 'meta-learning' (i.e. the overall learning from the country-level learning). | 81% | 14% | 5% | 2.0 | 8 |
| 2.17 | Develop instructional manuals on how to investigate and learn from patient safety incidents. | 80% | 20% | 0% | 1.0 | 8 |
| 2.18 | *Support improvement in incident reporting and learning through sharing exemplars.* | 76% | 19% | 5% | 1.0 | 8 |
| 2.19 | *Ability to share reported patient safety incidents/risks relevant to international learning.* | 76% | 19% | 5% | 2.0 | 9 |
| 2.20 | *Ability to analyse and make recommendations based on shared patient safety data to identify transferable learning from countries responding to patient safety risks.* | 76% | 24% | 0% | 2.0 | 8 |
| 2.21 | *Ability to trigger an alarm that deploys a notification to countries about serious identified patient safety risks.* | 76% | 19% | 5% | 2.0 | 8 |
| 2.22 | *Support improvements in collection of patient safety data through sharing exemplars.* | 76% | 19% | 5% | 2.0 | 8 |
| **Section 3A: Patient safety incidents relevant to international sharing and learning** | | | | | | |
| 3.01 | Incident involves a product or device that might be a major contributing factor to the incident. | 100% | 0% | 0% | 1.0 | 7 |
| 3.02 | Incidents where risk of severe harm or death is likely, should the same incident reoccur. | 100% | 0% | 0% | 1.5 | 8 |
| 3.03 | Incidents related to drug and equipment safety. | 100% | 0% | 0% | 1.5 | 8 |
| 3.04 | Incidents related to manufacturing and supply chain (e.g. contaminated vaccine/IV fluids/injections/drugs). | 95% | 5% | 0% | 1.0 | 8 |
| 3.05 | Incidents related to faulty medical devices (including IVDs) or equipment failure. | 95% | 5% | 0% | 1.0 | 8 |
| 3.06 | Incidents identified by multiple countries as common sources of unsafe care and therefore a potential priority to learn from and tackle collectively. | 95% | 5% | 0% | 2.0 | 8 |
| 3.07 | Patient safety risks that are new and/or have not been reported before in your country. | 93% | 7% | 0% | 1.5 | 8 |
| 3.08 | Patient safety risks related to diagnostic errors. | 93% | 7% | 0% | 1.5 | 7 |
| 3.09 | A near miss (events where no harm was done, but there could have been if people had done the same thing and, if allowed to go a stage further, it could have led to a death). | 87% | 7% | 7% | 1.0 | 7 |
| 3.10 | Incidents relevant to current international campaigns or challenges (e.g. the WHO patient safety challenges). | 86% | 14% | 0% | 2.0 | 8 |
| 3.11 | All adverse events (an incident that results in harm to a patient) (WHO, 2009). | 80% | 20% | 0% | 2.0 | 8 |
| 3.12 | *Ten times medication errors.* | 76% | 19% | 5% | 2.0 | 8 |
| 3.13 | *Tubing misconnections/misconnection errors (e.g. non-luer connected devices).* | 76% | 19% | 5% | 2.0 | 7 |
| 3.14 | *Nasogastric tube positioning errors/incidents.* | 76% | 19% | 5% | 2.0 | 7 |
| 3.15 | *Incidents that result from relatively novel contributory factors (e.g. pandemics, electricity outage, internet outage, civil unrest, war, funding issues, natural disasters, deliberate sabotage, criminal activity).* | 73% | 27% | 0% | 1.5 | 8 |
| 3.16 | *Any incident type that impacts paediatric patients.* | 73% | 27% | 0% | 1.5 | 7 |
| **Section 3B: Criteria used in the decision making of what should be shared internationally** | | | | | | |
| 3.17 | Risk of harming a large number of individuals in multiple countries if no intervention is taken. | 100% | 0% | 0% | 0.5 | 9 |
| 3.18 | Morbidity and mortality from patient safety risks/events (i.e. impact and severity). | 100% | 0% | 0% | 1.0 | 8 |
| 3.19 | The proposed patient safety solution needs international action (e.g. action from major pharmaceutical companies). | 100% | 0% | 0% | 1.15 | 8 |
| 3.20 | Identified patient safety risk(s) is/are relevant to more than one country facing similar clinical challenges (e.g. prevention and control of infectious disease). | 93% | 0% | 7% | 0.0 | 8 |
| 3.21 | Identified patient safety risk(s) is/are relevant to more than one country (e.g. supply of material/medicine/raw materials/devices originating in another country). | 87% | 7% | 7% | 1.0 | 8 |
| 3.22 | Ease of measurement of the identified patient safety risk(s) in multiple countries. | 87% | 7% | 7% | 1.5 | 8 |
| 3.23 | Stakeholders' interest on specific patient safety incidents/risks. | 80% | 20% | 0% | 1.0 | 7 |
| 3.24 | The availability of evidence to support unequivocal preventability. | 80% | 13% | 7% | 2.0 | 8 |
| **Section 4A: Key enablers to setting up an international PSLS** | | | | | | |
| 4.01 | Having a standardised international patient safety taxonomy. | 100% | 0% | 0% | 1.0 | 9 |
| 4.02 | Having the patient experience as a facilitator around which common definitions of harm and their priority for mitigation are set. | 100% | 0% | 0% | 1.5 | 7 |
| 4.03 | For users and/or contributors to the system to be able to access it in an easy, secure way. | 95% | 0% | 5% | 1.0 | 9 |
| 4.04 | Having buy-in from healthcare systems within each individual country. | 95% | 5% | 0% | 1.0 | 8 |
| 4.05 | The system is accessible for everyone to read the information that helps organisations and individuals learn (e.g. web page open for everyone). | 95% | 0% | 5% | 1.0 | 8 |
| 4.06 | To have representation from countries that are submitting data to the international system (e.g. national co-ordinators). | 95% | 5% | 0% | 2.0 | 8 |
| 4.07 | Having political will to generate and co-ordinate patient safety interventions of international concern. | 93% | 7% | 0% | 1.0 | 8 |
| 4.08 | Deploying resources/learning where they are needed most. | 93% | 7% | 0% | 1.0 | 8 |
| 4.09 | Encouraging and funding cross-jurisdictional studies of strategies designed and evaluated to understand better system functioning and implementation impact. | 93% | 7% | 0% | 1.0 | 7 |
| 4.10 | The benefits for those who are supposed to feed the information to the system are very clear. | 93% | 0% | 7% | 1.5 | 7 |
| 4.11 | Broad support from the clinicians and the societies that make up the international patient safety community. | 90% | 10% | 0% | 1.0 | 9 |
| 4.12 | Country-level will to learn with and from other countries. | 90% | 10% | 0% | 1.0 | 8 |
| 4.13 | Having buy-in from international organisations with a role and interest in patient safety. | 90% | 10% | 0% | 2.0 | 9 |
| 4.14 | Automation of as many functions as possible. | 90% | 5% | 5% | 2.0 | 9 |
| 4.15 | Having internationally comparative patient safety data. | 87% | 7% | 7% | 2.0 | 8 |
| 4.16 | Governments, agencies, and organisations have clear legislation, regulations and guidelines that are supportive of and encouraging of sharing of patient safety data relevant for international learning. | 86% | 5% | 10% | 2.0 | 8 |
| 4.17 | Involvement of important knowledge mobilisers (e.g. Institute for Healthcare Improvement, World Health Organization). | 86% | 14% | 0% | 2.0 | 8 |
| 4.18 | Existence of international standards/convention for sharing patient safety data. | 86% | 14% | 0% | 2.0 | 8 |
| 4.19 | An independent body/organisation responsible for operating and maintaining the international PSLS. | 81% | 19% | 0% | 1.0 | 7 |
| 4.20 | *Multiple input methods are available (e.g. online, phone, e-mail).* | 76% | 14% | 10% | 2.0 | 8 |
| 4.21 | *Password-controlled access to an online platform.* | 73% | 13% | 13% | 1.5 | 7 |
| 4.22 | *Funded by partner countries.* | 71% | 29% | 0% | 2.0 | 8 |
| 4.23 | *Access for input to the system should be limited and discussed nationally, with only one named responsible organisation in each country.* | 53% | 27% | 20% | 2.5 | 7 |
| **Section 4B: Key challenges/barriers to setting up an international PSLS** | | | | | | |
| 4.24 | Difficulty with funding this learning system even from participating countries. | 100% | 0% | 0% | 1.0 | 9 |
| 4.25 | Potential cost of establishing and maintaining the system. | 100% | 0% | 0% | 1.0 | 9 |
| 4.26 | Lack of a data/information governance strategy. | 100% | 0% | 0% | 1.5 | 8 |
| 4.27 | The 'what's in it for me' problem, i.e. to find the value proposition/business case in the various and varied jurisdictions. | 100% | 0% | 0% | 2.0 | 8 |
| 4.28 | Many countries have multiple reporting and learning organisations. | 93% | 0% | 7% | 1.5 | 9 |
| 4.29 | Limited resources in less developed countries. | 90% | 5% | 5% | 1.0 | 9 |
| 4.30 | Issues about national reputation and healthcare system reputation, which might create barriers. | 90% | 10% | 0% | 2.0 | 8 |
| 4.31 | Many countries are at very different maturity levels with respect to just/safety culture. | 87% | 7% | 7% | 2.0 | 8 |
| 4.32 | Finding the responsible international organisation to set up the system and relevant national organisations that will commit to co-operate. | 86% | 10% | 5% | 2.0 | 8 |
| 4.33 | At the international level, big cultural factors and different types of healthcare systems might present a challenge. | 86% | 10% | 5% | 2.0 | 8 |
| 4.34 | Concerns regarding privacy issues when sharing learning from investigations of patient safety incidents. | 86% | 10% | 5% | 2.0 | 8 |
| 4.35 | Having too many different bodies which are involved. | 81% | 19% | 0% | 1.0 | 7 |
| 4.36 | Lack of a sense of ownership in a system the end-user has not been involved in developing. | 81% | 14% | 5% | 2.0 | 8 |
| 4.37 | Lack of common format/approach to sharing learning about patient safety risks and/or efforts to mitigate/prevent harm to patients. | 81% | 14% | 5% | 2.0 | 8 |
| 4.38 | The information collected by this system will have limited utility if it is relying on system-centric versions of patient harm without explicitly including the patient perspective of harm. | 80% | 20% | 0% | 1.0 | 7 |

*IQR = interquartile range; IVD = in vitro diagnostic device; PSLS = patient safety learning system; WHO = World Health Organization. Shaded rows (light red) = statements not meeting the ≥80% consensus threshold.*

**Table S3: Newly generated statements from round one qualitative feedback**

Following the first round of the Delphi survey, free-text feedback from panellists was subjected to thematic analysis. This generated 33 new statements (organised by survey section) and a new subsection of 8 statements on criteria for international sharing (Section 3B), all of which were included in the second round of the survey.

| **Category** | **Newly Generated Statements** |
| --- | --- |
| **Statements generated from round one qualitative feedback, included in round two** | |
| **Purpose(s) of an international PSLS** | - Identify evidence-practice gaps that are occurring in similar contexts in other countries in order to develop interventions together. - Learn from patients and families about risk, harm, response, and remediation. - Bolster capability of those seeking to improve safety by benefitting from the shared learnings of those that have achieved improvements in safety in similar care contexts. - Learn with and from countries about efforts to improve patient safety in terms of what works and how. - Identify priorities for research and development to focus international efforts and resources where they are needed most. - Co-ordinate efforts of national patient safety organisations in collaboration with WHO. - Sharing affordable design-based interventions. |
| **Key features and functions of an international PSLS** | - Facilitate learning and support through a range of formats (e.g. instructional manuals, train-the-trainer workshops, webinars, onsite learning). - Develop a structured process for investigating patient safety risks identified by the international learning system. - Develop methodologies to evaluate practices to learn from patients and family members in all facets of patient safety, including prevention, response, recovery, and remediation. - Support the progression of countries towards a proactive and personalised clinical risk management approach (i.e. moving from a reactive to a proactive mindset). - To develop methodological approaches for assimilating a range of descriptive patient safety data to build a more complete understanding of safety within countries. |
| **Patient safety incidents relevant to international sharing and learning** | - Incidents where risk of severe harm or death is likely, should the same incident reoccur. - Incidents that result from relatively novel contributory factors (e.g. pandemics, electricity outage, internet outage, civil unrest, war, funding issues, natural disasters, deliberate sabotage, criminal activity). - Patient safety risks that are new and/or have not been reported before in your country. - Patient safety risks related to diagnostic errors. - Incidents related to drug and equipment safety. - Any incident type that impacts paediatric patients. - Incident involves a product or device that might be a major contributing factor to the incident. |
| **Key enabling factors for the setup of an international PSLS** | - Having political will to generate and co-ordinate patient safety interventions of international concern. - Having internationally comparative patient safety data. - Encouraging and funding cross-jurisdictional studies of strategies designed and evaluated to understand better system functioning and implementation impact. - Having a standardised international patient safety taxonomy. - Having the patient experience as a facilitator around which common definitions of harm and their priority for mitigation are set. - The benefits for those who are supposed to feed the information to the system are very clear. - Deploying resources/learning where they are needed most. |
| **Key challenges/barriers to the setup of an international PSLS** | - Many countries have multiple reporting and learning organisations. - Many countries are at very different maturity levels with respect to just/safety culture. - Difficulty with funding this learning system even from participating countries. - The 'what's in it for me' problem, i.e. to find the value proposition/business case in the various and varied jurisdictions. - Potential cost of establishing and maintaining the system. - The information collected by this system will have limited utility if it is relying on system-centric versions of patient harm without explicitly including the patient perspective of harm. - Lack of a data/information governance strategy. |
| **Additional statements generated from round one (new Section 3B criteria, included in round two)** | |
| **Criteria for deciding what patient safety incident/risk is of international concern for sharing and learning** | - Morbidity and mortality from patient safety risks/events (i.e. impact and severity). - Stakeholders' interest on specific patient safety incidents/risks. - Ease of measurement of the identified patient safety risk(s) in multiple countries. - The availability of evidence to support unequivocal preventability. - Identified patient safety risk(s) is/are relevant to more than one country (e.g. supply of material/medicine/raw materials/devices originating in another country). - Risk of harming a large number of individuals in multiple countries if no intervention is taken. - Identified patient safety risk(s) is/are relevant to more than one country facing similar clinical challenges (e.g. prevention and control of infectious disease). - The proposed patient safety solution needs international action (e.g. action from major pharmaceutical companies). |

*PSLS = patient safety learning system; WHO = World Health Organization.*
